# PUM1 represses CDKN1B translation and contributes to prostate cancer progression

**DOI:** 10.1101/2021.05.27.21257930

**Authors:** Xin Li, Jian Yang, Xia Chen, Dandan Cao, Eugene Yujun Xu

## Abstract

Posttranscriptional regulation of cancer gene expression programs plays a vital role in carcinogenesis; identifying the critical regulators of tumorigenesis and their molecular targets may provide novel strategies for cancer diagnosis and therapeutics. Highly conserved RNA binding protein PUM1 regulates mouse growth and cell proliferation, propelling us to examine its role in cancer. We found human *PUM1* is highly expressed in a diverse group of cancer, including prostate cancer; enhanced PUM1 expression is also correlated with reduced survival among prostate cancer patients. Detailed expression analysis in twenty prostate cancer tissues showed enhanced expression of *PUM1* at mRNA and protein levels. Knockdown of *PUM1* reduced prostate cancer cell proliferation and colony formation, and subcutaneous injection of *PUM1* knockdown cells led to reduced tumor size. Downregulation of PUM1 in prostate cancer cells consistently elevated CDKN1B protein expression through increased translation but did not impact its mRNA level, while overexpression of PUM1 reduced CDKN1B protein level. Our finding established a critical role of PUM1 mediated translational control, particularly the PUM1-CDKN1B axis, in prostate cancer cell growth and tumorigenesis. We proposed that PUM1-CDKN1B regulatory axis may represent a novel mechanism for the loss of CDKN1B protein expression in diverse cancers and could be potential targets for therapeutics development.

## Introduction

Prostate cancer is the most common cancer and one of the leading causes of cancer death in men. Overexpression of RBPs (RNA Binding Proteins) in prostate cancer (PCa) supports that posttranscriptional control plays a vital role in prostate carcinogenesis ^[1]^. Only a few posttranscriptional regulators have been identified, and even fewer are mechanistically understood ^[2,3]^.

Human homologs of a significant proportion of mouse growth genes are also essential for tumorigenesis, either as oncogenes or tumor suppressors. Many of the growth regulators or tumorigenesis regulators are transcriptional regulators ^[4]^, the role of posttranscriptional regulation has begun to be appreciated ^[2]^. Characterization of conserved RNA binding proteins in mammalian growth and tumorigenesis could help identify novel regulators and pathways important for cancer diagnosis and therapeutics development.

One of the highly conserved eukaryotic RNA-binding proteins, the PUF family (Pumilio and FBF), functions by binding to Pumilio Binding Elements (PBE) on the 3′UTR of their target mRNAs and play critical roles in cell fate decision and differentiation ^[2,5-7]^. Recent genetic studies on members of the mammalian PUF family, *Pum1* and *Pum2*, uncovered roles of PUM-mediated posttranscriptional regulation in growth, reproductive and neuronal development as well as diseases ^[8-16]^. *Pum* mutant mice have reduced body weight and size, resulting from enhanced cell cycle inhibitor expression, *Cdkn1b*, and reduced cell proliferation, raising the possibility that PUM-CDKN1B regulatory pathway may also be important in human growth and cell proliferation ^[15]^.

The p27Kip1 (*Cdkn1b*) gene is a member of the Cip/Kip family of cyclin-dependent kinase (CDK) inhibitors that function to negatively control cell cycle progression by association with cyclin-dependent kinase 2 (CDK2) and cyclin E complexes to inhibit the transition from G1 to S phase ^[17]^. Previous work has demonstrated that *CDKN1B* is a haploinsufficient tumor suppressor whose protein level has to be fine-tuned for its optimal function, and CDKN1B protein level may have prognostic and therapeutic implications ^[17-19]^. While impaired synthesis, accelerated degradation, and mislocalization of CDKN1B have been studied ^[19]^, regulation of CDKN1B protein expression via translation in cancer has not been explored, and whether there exist key translational regulators remained unknown. We propose that *Pum1* may exert control on human cell growth, in particular cancer cell growth, by repressing *Cdkn1b* expression ^[15]^.

Association of PUM with cancer cell growth has been suggested from studies on cancer cell lines ^[20-22]^, examination of PUM expression in human tumorigenesis and characterization of PUM mediated translation control directly in human cancer cells and the tumor is needed to reveal the extent and mechanism of PUM-mediated regulation during human tumorigenesis. Here, we report increased expression of PUM1 in various cancers from an extensive collection of human cancer datasets and twenty prostate tumor samples we collected from the clinic. We uncovered a role of PUM1-mediated translational repression of CDKN1B in prostate cancer cell proliferation and tumor formation. Our data underscore the importance of the PUM1-CDKN1B regulatory axis in human carcinogenesis and its potential for developing novel strategies in cancer diagnosis and therapeutics.

## Material and Methods

### Patients and Tissue Samples

All human studies were approved by the Institutional Ethics Committee of Nanjing Medical University and performed after obtaining written informed consent. Twenty patients (mean age 70.5 years, range 61-76 years) underwent radical prostatectomy for prostate adenocarcinoma at the Second Affiliated Hospital of Nanjing Medical University. A summary of clinic and pathology parameters for the study cohort is included in Supplemental Table II. For prostate cancer samples, the predominant tumor nodule and matched normal tissue were identified by histopathologists on fresh-frozen sections. Only foci with >80% purity of cancer cells were collected. Each prostate cancer and matched normal tissue were cut into three pieces. Among them, two parts were immediately frozen in liquid nitrogen and stored at −80°C for later protein and RNA extraction, one piece of tissue was fixed in 10% neutral buffered formalin for 24 hours. The procedure for this research conforms to the provisions of the Declaration of Helsinki.

### Immunohistochemistry

The histologic specimens were fixed in 10% formalin and routinely processed for paraffin embedding. Histologic sections, 5-mm thick, were stained with hematoxylin-eosin and reviewed by two pathologists to define the prostate cancer and matched normal tissues. Immunohistochemistry was performed on tumor paraffin sections after antigen retrieval using antibodies directed against PUM1 (Abcam, ab92545), according to the protocol described previously ^[15]^. Specificity of PUM1 antibody has been confirmed using tissues from *Pum1* knockout mice ^[15]^.

### Western Blot

Total protein was collected using the RIPA (Beyotime, P0013B) and protease inhibitor cocktail (Roche, Basel). Standard Western blotting procedure ^[15]^ was followed with PVDF membrane (Bio-Rad) used for protein transfer. Detection of HRP conjugated secondary antibody was performed with ECL (PerkinElmer). The antibodies used were as follows: Rabbit anti-PUM1 (Abcam, ab92545), mouse anti-ACTIN (SIGMA, A1978), rabbit anti-CDKN1B (CST, #2552P), mouse anti-GAPDH (Protein tech, 60004-1-Ig), Mouse anti-Tubulin (Santa Cruz, sc-8035). The band intensity of specific proteins was quantified after normalization with that of β-ACTIN or GAPDH.

### RNA Extraction and RT-PCR Analysis

RNA was extracted by Trizol from tissues or cells stored at −80°C and reverse transcribed for amplification of *PUM1* cDNA. RT-qPCR analysis was performed using gene-specific sets of primers (Supplementary Table SI). The gene expression analysis was detected by ABI BioSystem StepOne plus. The gene expression level was quantified relative to the expression of *β-ACTIN*, and the specificity of PCR products was confirmed by melting curve analysis. Each reaction filled up to an end volume of 20 μL containing two μl template cDNA, ten μl SYBR Premix Ex Taq buffer (TaKaRa, RR820A), 0.4 μl ROX Reference Dye, eight pmol of each primer, and six μl ddH2O and was carried out in a standard 96-well plate. The cycling conditions consisted of an initial incubation at 95 °C for 3 min, followed by 45 cycles of 94°C for 30 s, 60°C for 30 s, and 72°C for 30 s. A final incubation terminated the reaction at 95°C for 15 s, 60°C for 30 s, and 95°C for 15 s. The expression level was calculated by the 2-ΔΔCt method to compare the relative expression.

### Luciferase reporter assay

*CDKN1B* 3′UTR was subcloned into psiCHECK-2 vector (Promega) using XhoI and PmeI restriction enzymes (New England Biolabs). Wild-type PBE sequences 5′-TGTATATA-3′ was mutant to 5′-acaATATA-3′ as previous report ^[12]^. Cells were co-transfected with pCMV6-hPUM1 and luciferase reporter plasmids, containing fragments or full-length 3’UTRs. After 48 hours, cells were washed with PBS and lysed in Passive Lysis Buffer (Promega, Chroma-Glo™ Luciferase Assay System). 20μl of each lysate was analyzed using the Dual-Luciferase Reporter Assay System (Promega, Chroma-Glo™ Luciferase Assay System) in a 96 Microplate luminometer (BioTek, USA).

### Sucrose gradient polysome fractionation

2×10^7^ DU145 cells were collected, washed with PBS, and homogenized in 0.5mL of MCB buffer. The lysate was centrifuged at 1300g at 4°C for 10 min. The supernatant was applied onto the top of a 15–55% (W/W) linear sucrose gradient made by the Density Gradient Fractionation System (Teledyne ISCO Inc.). The gradient was centrifuged at 150000g for three hours (Beckman, USA). Fractions were collected and used for RNA extraction and analysis.

### Colony formation

DU145 cells were generated by hPUM1 shRNA or scramble control lentiviral infection followed by puromycin selection. Cells were then trypsinized and counted by a hemocytometer. In total, 1 × 10^4^ *PUM1* knockdown or overexpressing DU-145 cells were seeded in complete growth media and allowed to grow for 14–21 days until visible colonies formed. Colonies were stained with 0.25 % crystal violet in ddH2O, washed with PBS twice, and air-dried.

### Xenograft model in vivo

Six-week-old male nude mice (BALB/c, Charles River) were used for the xenograft experiments. Cancer cells were trypsinized and harvested in PBS, then a total volume of 0.1 ml PBS was subcutaneously injected into the inguinal regions. DU-145 cells (5×10^5^) transduced with sh-PUM1KD1 or Sh-Con were subcutaneously injected into the left and right inguinal regions of nude mice, respectively and the nude mice were monitored for 49 days. Tumor sizes were measured twice a week, starting at two weeks after cell injection using Vernier caliper. The animal study protocol was reviewed and approved by the institutional animal care and use committee of Nanjing Medical University.

### Statistical analysis

All experiments were repeated at least three times. Statistical significance between two groups of data was evaluated by Student’s t-test (two-tailed) comparison using GraphPad Prism software 7.

## Results

### PUM1 expression is upregulated in various cancers, and overexpression of PUM1 is associated with poor prognosis in prostate cancer patients

To determine whether PUM1 has a role in tumorigenesis, we first examined PUM1 expression in the TCGA database and found that PUM1 is widely expressed in diverse cancers, including prostate cancer (Fig. 1A). Given the reported correlation of loss of CDKN1B expression with prostate cancer survival ^[18]^, we hence investigated the role of PUM1 expression in prostate cancer. By focusing on 101 prostate samples from the TCGA database ^[23]^, we found that *PUM1* mRNA was significantly higher in prostate carcinoma than that in benign tissues (P<0.05) (Fig. 1B). Compared with the normal prostate gland, a substantially higher level of PUM1 protein in prostate carcinoma was observed by immune-histochemical staining assays (Fig. 1C). Furthermore, the Kaplan-Meier curves indicated a significant correlation of PUM1 protein upregulation with poor patient survival (Fig. 1D). Hence, our analysis of TCGA cohorts suggested that PUM1 upregulation is associated with tumorigenicity, particularly prostate cancer, and poor outcomes in PCa patients.

**Figure 1.**
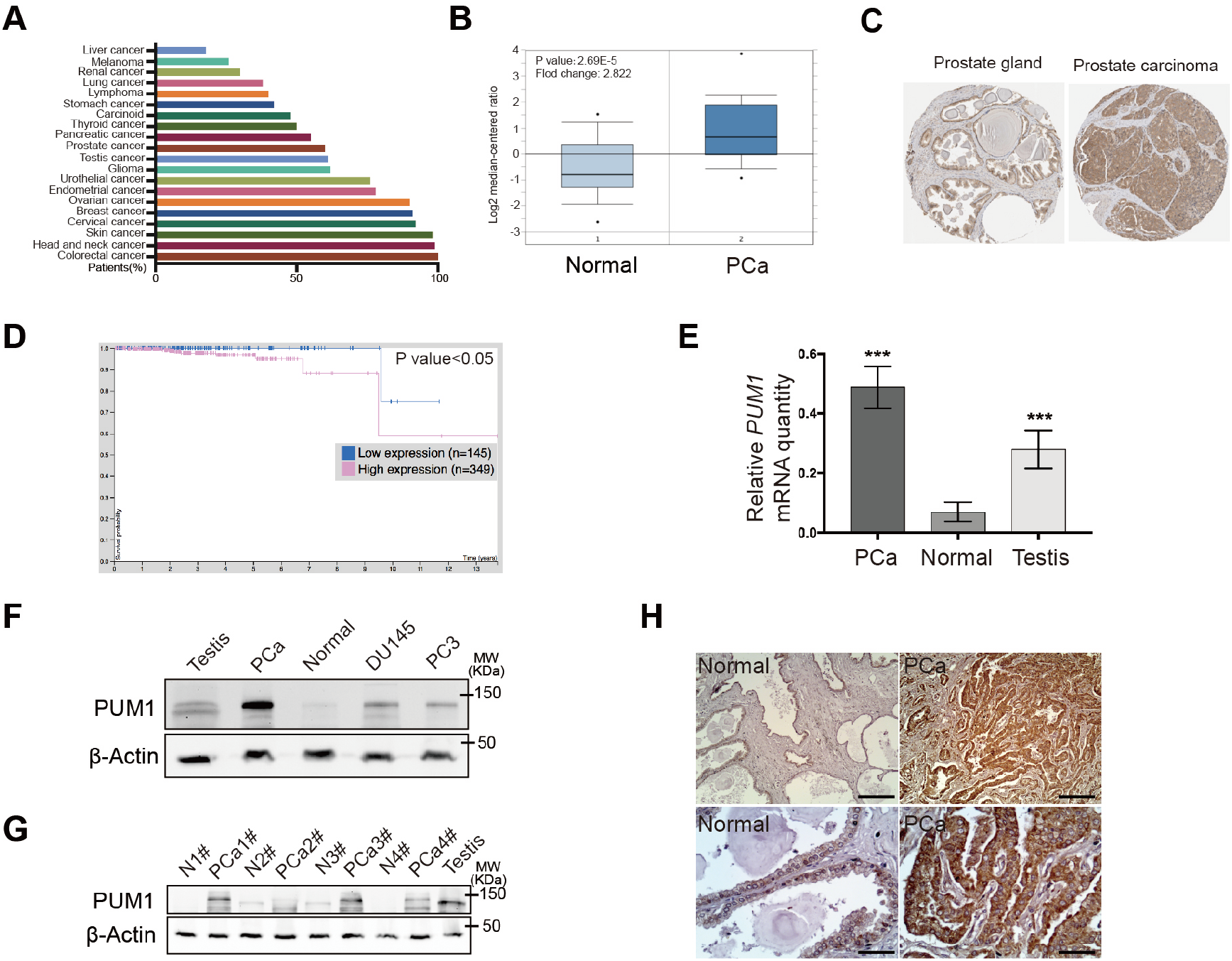
Overexpression of PUM1 is associated with poor prognosis in patients with PCa. (A). Graph showing percentage of patients staining positive for PUM1 protein among TCGA cancer types using validated anti-PUM1 antibody from Human Protein Atlas. (B). The box plot comparing *PUM1* expression in the normal prostate gland (Normal) (n=29) and prostate carcinoma (PCa) (n=72) was derived from the Oncomine database (https://www.oncomine.org/). (C). The expression of PUM1 in the normal prostate gland and prostate adenocarcinoma specimens. Representative images were taken from the Human Protein Atlas database. (D). The Human Protein Atlas survival analysis for low and high expression levels of PUM1 on 494 prostate cancer patients with death outcome. (E). RT-qPCR examined the expression of PUM1 mRNA in prostate cancer tissue (PCa) and normal prostate tissues (Normal). Human testis (Testis) is a positive control since *PUM1* is highly expressed in the testis. ***P < 0.001. (F). PUM1 protein is increased in prostate cancer cell lines (DU145 and PC3) and tumor tissue from one cancer patient. (G) PUM1 protein level was detected in adjacent normal tissue (N) and prostate carcinoma tissue (PCa) of different cases. (H). Immunohistochemistry of PUM1 protein in prostate carcinoma (PCa) and normal prostate gland (Normal). Scale bar: 200μm (up panel) and 50μm (low panel).

To further verify the correlation between PUM1 expression and PCa, we collected and analyzed PCa specimens from 20 prostate carcinoma patients in the clinic (Supplemental Table II). RT-qPCR from total RNA obtained from PCa tissue and matched normal tissue confirmed the upregulation of *PUM1* at the mRNA level in most of the patients examined (Fig. 1E). Western blot analysis was performed on extracts obtained from the neoplastic tissue and from the gland′s contralateral part. We found that PUM1 protein was consistently upregulated in the neoplastic tissues, mostly basal cells (Fig. 1F and 1G). PUM1 protein was also detected in two types of prostate cancer cells (DU145 and PC3) (Fig. 1F). To further determine the localization of PUM1 protein in the prostate cancer tissue, we performed immunohistochemistry analysis on tissues of the same group of patients. PUM1 was expressed at low levels in the epithelial cells of matched normal prostate glands. By contrast, all epithelial cells of neoplastic glands were strongly positive for PUM1(Fig. 1H), which confirmed that expression of PUM1 is indeed elevated in the neoplastic phenotype of prostate epithelial cells.

### Downregulation of PUM1 in prostate cancer cells decreased proliferation and induced apoptosis

To understand the role of PUM1 in prostate carcinogenesis, we first interrogated the role of PUM1 in prostate cancer cell lines. We chose two commonly used prostate cancer cell lines representing different metastatic potentials of prostatic adenocarcinoma for our experiments, with PC3 from grade IV adenocarcinoma with high metastatic potential and DU145 from prostate carcinoma with moderated metastatic potential. Two *PUM1* small hairpin knockdown shRNAs were designed against different parts of the *PUM1* sequence and transfected into DU145 and PC3 cells. We found that *PUM1* mRNAs were significantly reduced in DU145 and PC3 cells transduced with the *PUM1* knockdown shRNA, confirming specific and efficient repression of PUM1 (Fig. 2A). Western blot analysis confirmed the reduction of PUM1 protein in both DU145 and PC3 cells containing either knockdown construct (Fig. 2B). Consistent with the previous report in mouse cells ^[12]^, CCK8 assays indicated that the downregulation of *PUM1* causes a decrease in the proliferation rate of DU145 cells (Fig. 2C). Growth inhibition by *PUM1* knockdown was also confirmed in PC3 cells (Fig. 2D). Annexin V/propidium iodide (PI) assay showed the population of apoptotic cells increased 1.41- and 1.35-fold in DU145, and 2.14- and 2.26-fold in PC3 when PUM1 was knocked down in these cells (Fig. 2E and 2F). Moreover, cleaved Caspase-3 and cleaved Caspase-9 were also increased in both cell lines upon *PUM1* knockdown (Fig. 2G). The reduced cell growth was due to inhibition of G1/S transition: *PUM1* knockdown significantly reduced DU145 cells in the S phase as assessed by incorporation of Edu (Fig. 2H) and DNA content (Fig. 2I). Together with our data demonstrated that an elevated PUM1 level is critical for the increased growth rate of prostate cancer cells.

**Figure 2.**
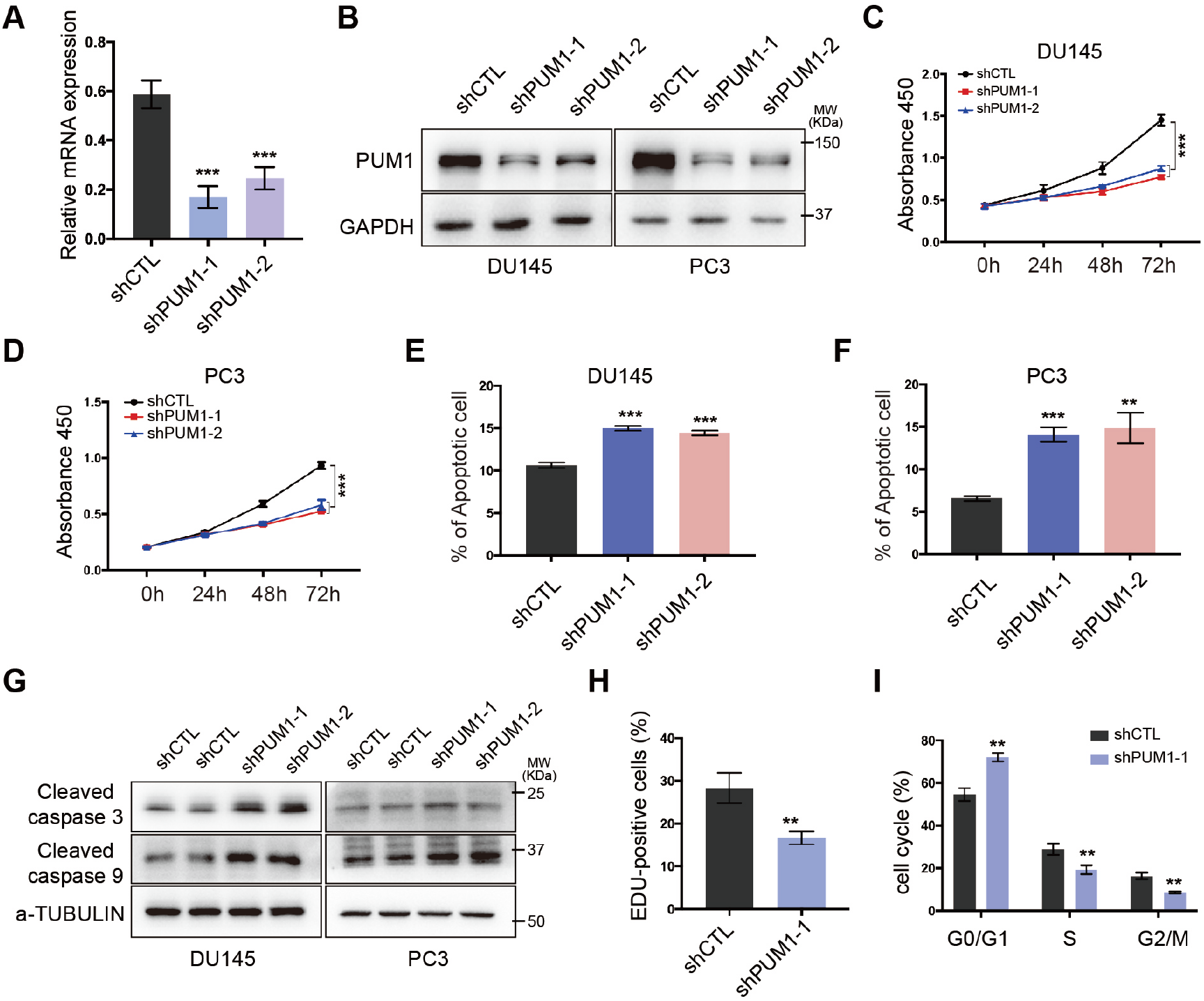
*PUM1* knockdown reduces PCa cell proliferation and survival. (A). RT-qPCR analysis of *PUM1* mRNA levels as normalized to *β-Actin* from DU145 cells. Two shRNAs (shPUM1-1 and shPUM1-2) target different *PUM1* transcript and non-targeting shRNA (shCTL) as a control. (B). Immunoblots showing the effect of shRNA-mediated *PUM1* knockdown in DU145 and PC3 cells. (C). CCK8 assay on DU-145 cells transfected with the shCTL or with *PUM1* shRNAs as described in (A). ***P < 0.001. (D). PC3 cells showed reduced growth after transfected with shRNAs knockdown construct. ***P < 0.001. (E) and (F). Apoptosis assay using flow cytometry after staining with annexin V-FITC/PI. ***P < 0.001, **P < 0.001. (G). Western blotting analysis of apoptosis-related proteins in PC3 and DU145 cells with *PUM1* knockdown. (H). EdU labeling of *PUM1* knockdown DU145 cells revealed significantly reduced proliferation in comparison with control cells. (I). Cell cycle analysis revealed increased G1 and decreased S and G2/M stage cells in *PUM1* Knockdown DU145 cells. **P <0.01.

### Overexpression of *PUM1* promotes prostate cancer cell proliferation and colony formation

Since PUM1 level is positively correlated with prostate cancer cells’ proliferation rate, we asked if overexpression of *PUM1* could further promote the proliferation of prostate cancer cells. The expression levels of PUM1 in DU145 cells were manipulated by transfection of *PUM1* knockdown construct and/or retroviral vector for *PUM1* (Fig. 3A). *PUM1* overexpression significantly increased the growth rate of DU145 (Fig. 3B). When PUM1 level was restored by retrovirus vector carrying *PUM1* gene, the growth rate of *PUM1*-knockdown cells became comparable to that of control cells (shCTL + Empty retroviral vector), supporting *PUM1* knockdown being responsible for reduced proliferation (Fig. 3B).

**Figure 3.**
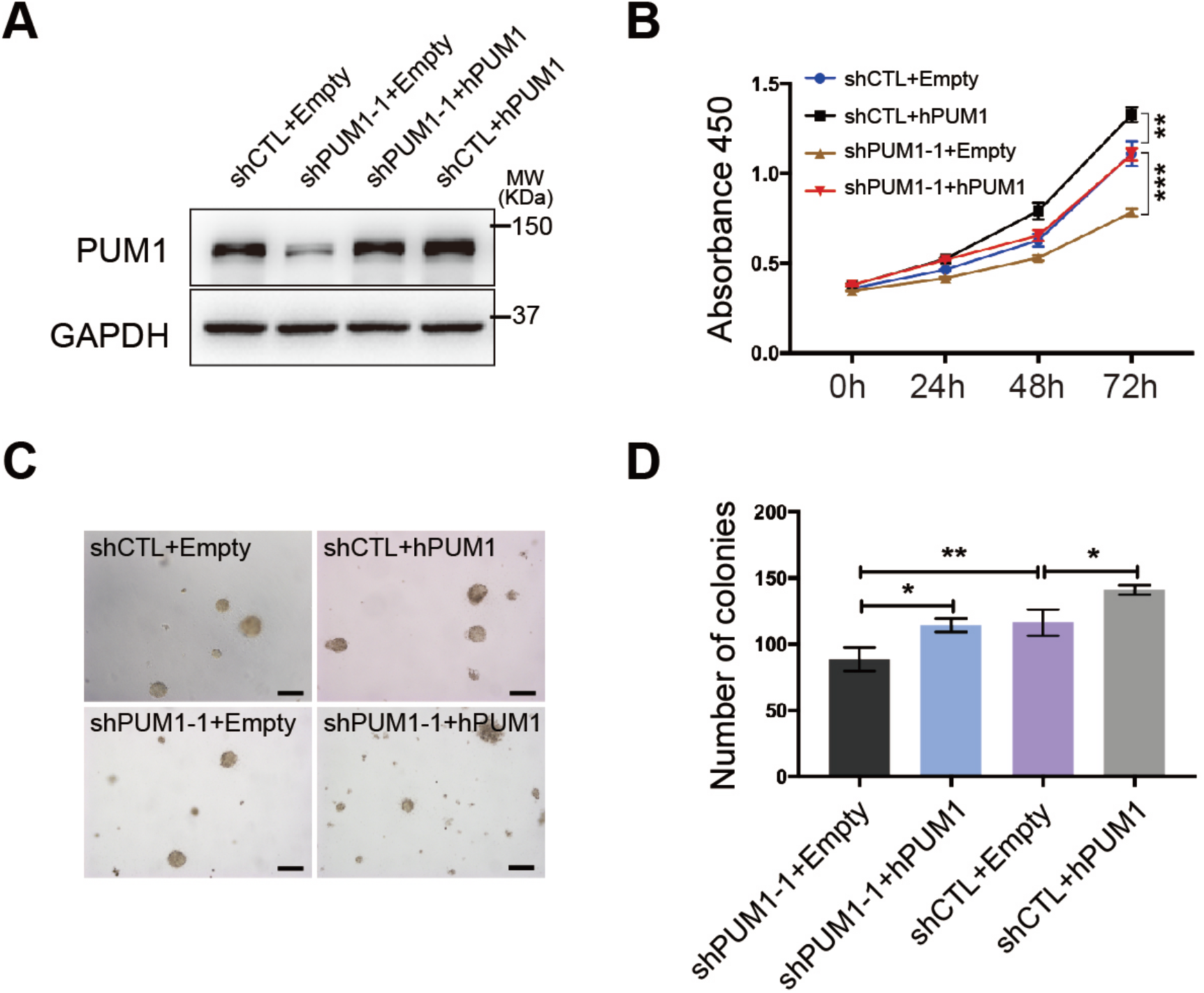
Overexpression of *PUM1* promotes prostate cancer cell proliferation and colony formation. (A). Western blot analysis of PUM1 protein expression from cells overexpressing *PUM1* or *PUM1* and shPUM1 at the same time. (B). Cell proliferation assay on *PUM1* overexpression and *PUM1* knockdown. (C). Colony assays were performed on the effect of overexpression of *PUM1* or knockdown of *PUM1* on colony number and size. (D). Colony number was counted for cells containing *PUM1* overexpressing plasmids or knockdown plasmid, or both. *P < 0.05, **p < 0.01.

We next asked the role of PUM1 in cell transformation by measuring the effect of PUM1 expression levels on the anchorage-independent growth of DU145 cells in soft agar assay. The total number of colonies was significantly reduced with *PUM1*-knockdown, while overexpression of *PUM1* led to a substantially higher number of colonies (Fig. 3C and 3D). Overexpressing *PUM1* in the cells with *PUM1*-knockdown rescued the ability of DU145 cells to form a comparable number of colonies, consistent with its rescue effect in cell proliferation. The positive correlation between PUM1 expression level and anchorage-independent growth further supports our hypothesis that PUM1 plays a critical role in prostate carcinogenesis.

### Reduced PUM1 expression represses prostate tumorigenesis *in vivo*

To test our hypothesis that PUM1 is essential for the growth of prostate cancer cells, we examined the effect of *PUM1*-knockdown on the tumorigenic capacity of DU145 cells in nude mice. DU145 cells transduced with an shRNA for *PUM1* (sh-PUM1) or a control shRNA (ShCTL) were injected into the left and right inguinal regions of the same nude mice (Fig. 4A and 4B). By both tumor weight and volumes, knockdown of *PUM1* significantly reduced the tumor growth (Fig. 4C and 4G). These results confirm the oncogenic activity of *PUM1* in vivo and suggest that *PUM1* may represent a potential target of therapeutic treatment for prostate cancer. Further exploration and investigation into PUM1’s regulatory mechanism could provide insight into such an approach’s feasibility.

**Figure 4.**
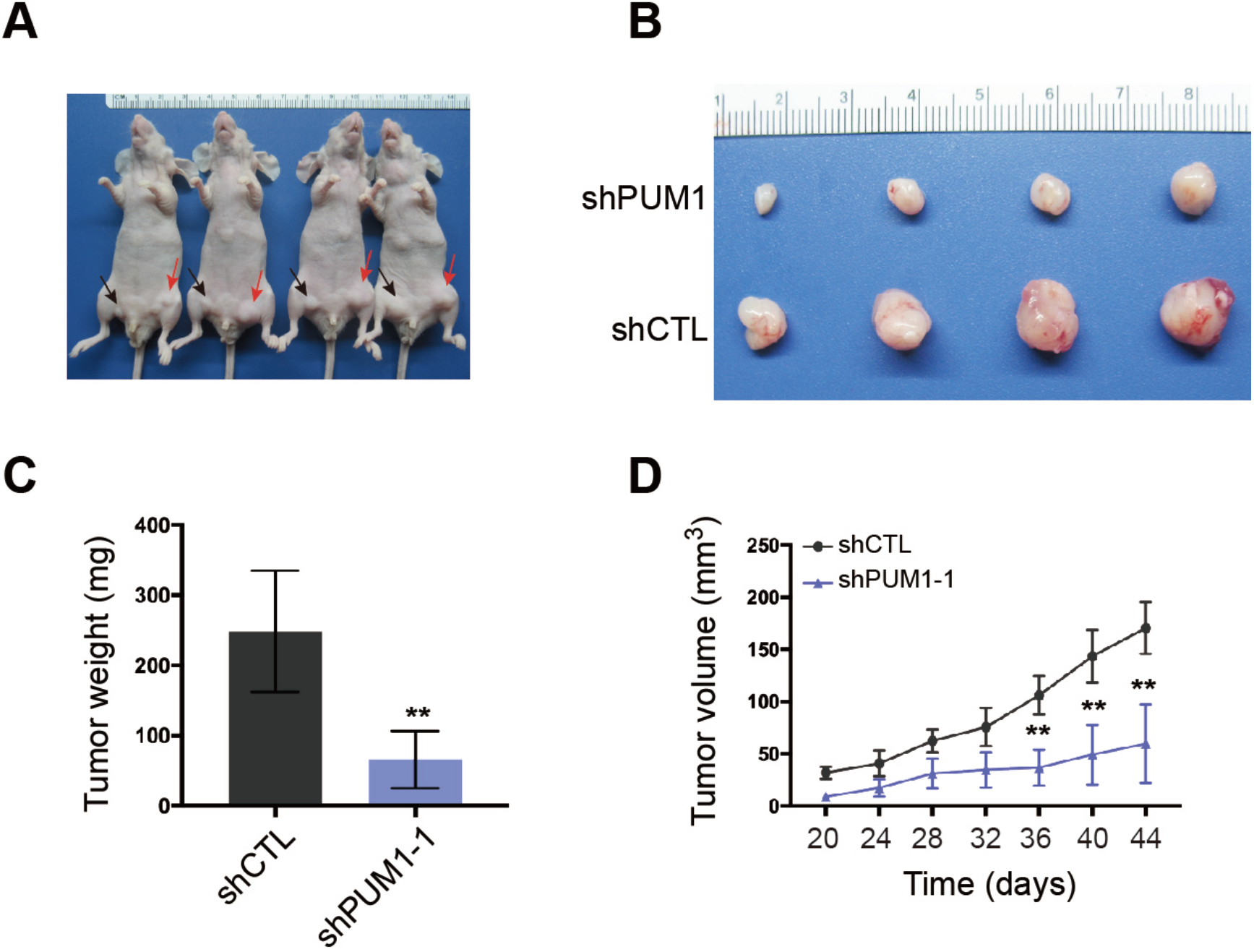
Knockdown *PUM1* reduces tumorigenesis of DU145 cells in vivo. (A and B). Representative images of xenografted nude mice (A) and tumors formed from *PUM1* knockdown cells and control cells (B). The black arrowhead indicates *PUM1* knockdown cells, and the red arrow indicates control cells. (C). Tumors from each group of mice were dissected and weighed for comparison of *PUM1* knockdown and shCTL. Histogram showing average tumor weight (N=4) was significantly lower for *PUM1* knockdown cells by comparison with control cells, **P<0.01. (D). Tumor volumes from each group of mice were measured on the indicated days. **p < 0.01.

### PUM1 represses the translation of tumor suppressor CDKN1B via binding to its PBE in prostate cancer cell

Genetic analyses established that *Cdkn1b* is a tumor suppressor in the prostate controlling prostatic epithelium growth ^[19]^. According to TCGA data, *CDKN1B* expression was observed to be downregulated in prostate cancer compared to normal tissues (Fig. 5A). Base on previous reports, we wondered whether PUM1 is involved in tumorigenesis via regulating CDKN1B expression. We then evaluated the correlation between PUM1 and CDKN1B gene expression with the TCGA-based GEPIA tool, and the plots showed that PUM1 and CDKN1B expression is correlated in PCa with Pearson correlation coefficient being 0.63 (Fig. 5B). RT-qPCR showed *CDKN1B* mRNA was unaffected in both DU145 and PC3 cells with *PUM1*-knockdown (Fig. 5C). To determine the molecular mechanism by which PUM1 contributed to cell proliferation and progression of prostate cancer, we analyzed the protein expression change of cell cycle regulators after inhibiting PUM1 expression. While other target cell cycle regulators exhibited variable changes or no changes in PUM1 knockdown cells, the protein expression of CDKN1B consistently increased upon knockdown of *PUM1* in both PC3 and DU145 cell lines, supporting CDKN1B as a major cell cycle downstream target of PUM1 protein (Fig. 5D and 5E).

**Figure 5.**
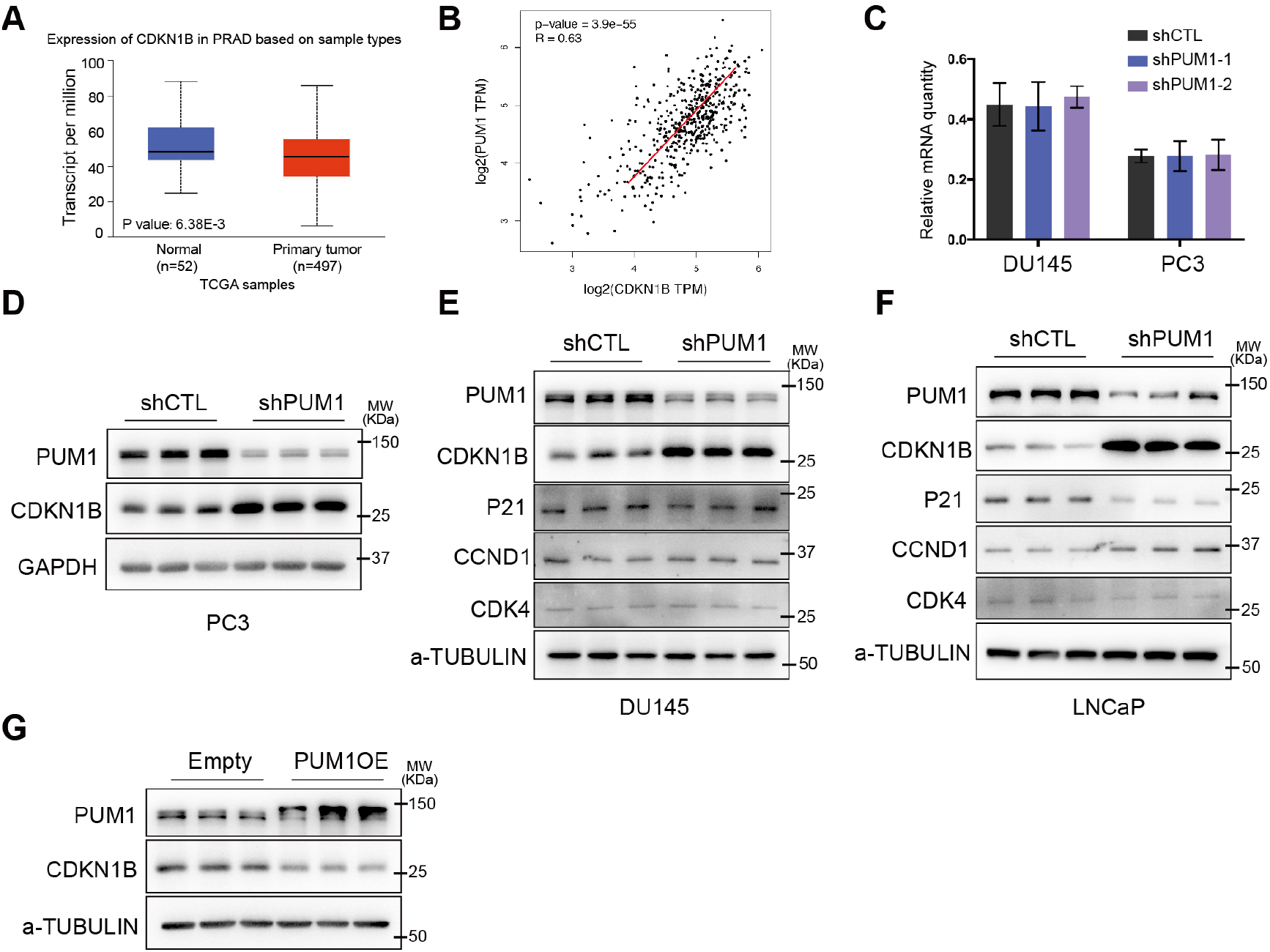
PUM1 represses the translation of CDKN1B mRNA by binding to the PBE motif on its 3’UTR. (A). Boxplots showed the expression of *CDKN1B* between normal prostate (n=52) and prostate carcinoma (n=497). Data were obtained from the GEPIA database (http://gepia.cancer-pku.cn/). (B). The correlation analysis of *PUM1* and *CDKN1B* in prostate cancer using the GEPIA tool showed that the mRNA expression level of the two genes was correlated to some extent. (C). RT-qPCR analysis of *CDKN1B* mRNA levels in DU145 and PC3 cells with *PUM1* knockdown. (D, E, and F). Western blot analyses of cell cycle proteins expression change from PC3, DU145, and LNCaP cells after 72 hours transfection with shPUM1-1 or shCTL. (G). Western blot analysis of CDKN1B protein level from DU145 cells transfected with human *PUM1* (PUM1OE) or empty plasmids.

To further determine if PUM1-mediated repression of CDKN1B is specific to androgen-insensitive DU145 and PC3 cell lines or general to all different types of prostate cancer cells, we chose LNCap cell line, which differ from DU145 and PC3 in that LNCap not only is from prostate carcinoma with low metastatic potential but also expresses androgen receptor and is androgen-sensitive. Knockdown of *PUM1* led to a significant reduction of PUM1 expression. The examination of cell cycle regulators showed a various degree of protein expression changes, CDKN1B exhibited the highest increase in protein expression, supporting PUM1-mediated repression of CDKN1B may be a general mechanism among prostate cancer cells (Fig. 5F). CCND1 protein was slightly increased while p21 protein is slightly decreased (Fig. 5F). These results suggested that PUM1-CDKN1B axis may represent a major regulatory axis in both AR-positive and -negative PC cells.

Given that CDKN1B protein but not RNA increased in the *PUM1* knockdown cells, we performed RNA immunoprecipitation (RIP) and dual luciferase assay to test if PUM1 directly regulates the translation of CDKN1B via binding to the two PBEs of its 3′UTR in prostate cancer cells. In the RNA immunoprecipitation experiment, we found that the pull-down fraction of PUM1 protein was significantly enriched with *CDKN1B* mRNAs, consistent with direct binding of PUM1 (Fig. 6A and 6B). Thus, *CDKN1B* mRNA is associated with PUM1 protein in DU145 and PC3 cells. Next, we constructed a luciferase reporter construct containing wildtype 3′UTR or mutant 3′UTR of *CDKN1B* mRNA with both PBE sites mutated (Fig. 6C). While PUM1 overexpression repressed expression of the reporter expression with wildtype *CDKN1B* 3′UTR, mutations on PBEs negated the repressive effect of PUM1 on the reporter (Fig. 6C). These results supported that PBE sites on the 3′UTR of *CDKN1B* are critical for PUM1 mediated translational repression in prostate cancer cells.

**Figure 6.**
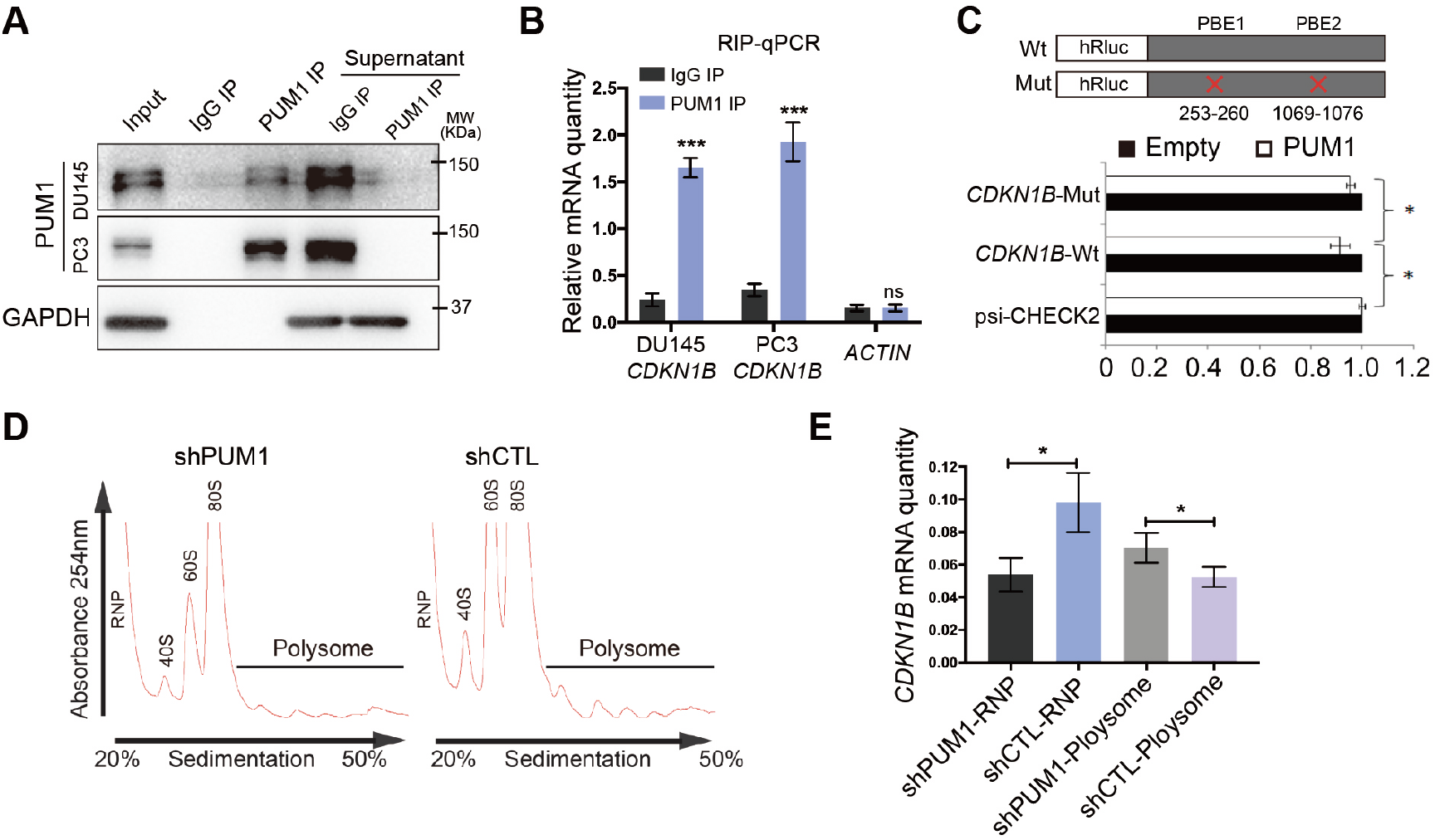
PUM1 represses *CDKN1B* translation via binding to the 3’UTR of *CDKN1B* mRNA. (A). RNA immunoprecipitation by PUM1 antibody from DU145 and PC3 cell lysates showed PUM1 protein could be specifically pull-down. (B). The RT-qPCR result using PUM1 immune-precipitates versus IgG precipitates showed that *CDKN1B* mRNA was highly enriched from PUM1 IP compared to IgG precipitates. (C). Diagram of wildtype and mutant *CDKN1B* 3’UTR containing two PUF binding elements (PBE) cloned in psiCHECK2 vector. Normalized luciferase activity expressed by the Luc-*CDKN1B* 3′-UTR constructs, co-transfected with the pCMV6-*hPUM1* vs. pCMV6 (Control) in 293T cells. *P < 0.05. (D) Polysome profiles from polysome fractionation experiments of DU145 cells lysate of *PUM1* knockdown (shPUM1) and control. (E) RT-qPCR detected the distribution of CDKN1B mRNA in free RNP or polysome fractions. *CDKN1B* mRNAs are significantly increased. *P < 0.05.

To test if PUM1 regulates the translation CDKN1B expression in prostate cancer cells, we performed a polysome fractionation experiment in DU145 cells transfected with or without a *PUM1* knockdown construct. *PUM1* knockdown significantly enriched CDKN1B mRNA in actively translating fraction, polysome fraction (Fig. 6D and 6E), indicating CDKN1B protein translation is promoted by *PUM1* knockdown. Together, these results supported our hypothesis that increased PUM1 expression may contribute to the growth of prostate cancer cells via translational repression of cell cycle inhibitor—CDKN1B and PUM1-CDKN1B regulatory axis may be an important regulatory mechanism in cancer cell proliferation.

## Discussion

In this study, we found PUM1 is frequently upregulated in human prostate carcinoma, and downregulation of PUM1 expression reduced PCa cell proliferation and survival. Our data from human cancer cells unveiled a conserved translational regulation of cell cycle regulators by PUM1 in tumorigenesis and prostate cancer progression. Overexpression of *PUM1* contributes to carcinogenesis by repressing the expression of negative cell cycle regulator CDKN1B, unveiling a novel mechanism for the loss of CDKN1B protein expression in tumorigenesis.

In cancer, the genetic control of the cell cycle is altered, resulting in unchecked growth and massive cell proliferation. The cell cycle-dependent kinase N1B (CDKN1B) is a negative regulator of the cell cycle and a tumor suppressor. *CDKN1B* is altered in 1.85% of all cancers with breast invasive ductal carcinoma, prostate adenocarcinoma, lung adenocarcinoma, colon adenocarcinoma, and testicular mixed germ cell tumor having the greatest prevalence of alterations ^[24]^. Decrease, but not a complete loss of CDKN1B protein activity could stimulate tumorigenesis and is proposed to be an essential step in the development and maintenance of malignant prostatic epithelial cell phenotype ^[19,25]^. Indeed downregulation of CDKN1B is found in most human prostate cancer^[25]^ and likely in most human tumors ^[26]^. Posttranscriptional regulation of CDKN1B appears to be a primary mechanism for CDKN1B downregulation during tumorigenesis ^[19,26],[27]^, Increased CDKN1B protein degradation via skp2 cks1or protein mislocalization through posttranslational modification were shown to control CDKN1B protein abundance and its tumorigenic function. While RNA binding protein HuD was implicated in reduced expression of CDKN1B in pancreatic cancer ^[28]^, but it is not known if translational regulation of CDKN1B abundance level is important for prostate cancer. Our finding that CDKN1B level is regulated at the translational level in prostate cancer cells by PUM1 protein revealed a novel mechanism regulating CDKN1B protein expression in prostate cancer, supporting the important roles of translational control in tumorigenesis.

Previously, it was reported the PUM1-mediated E2F3 post-transcription control may be necessary for the growth of cancer cell lines ^[21]^, and recently, PUM has been shown to be important for myeloid leukemia cell growth as well as hematopoietic stem cell growth ^[22]^. Abnormal expression of PUM proteins was also shown to cause genomic instability ^[29,30]^. However, it was unknown if PUM1 regulates tumorigenesis of solid tumors and to what extent PUM1-CDKN1B regulatory axis is important for cancer cell proliferation and tumor growth. Our findings implicate that PUM1 is a growth regulator in human prostate cancer and potentially a number of other cancers where PUM1 is highly expressed. We identified CDKN1B as the main target of PUM1 in a range of prostate cancer cells with features of different aggressiveness and hormone dependency, including both androgen-insensitive and androgen-sensitive cells. Consistent with the downregulation of CDKN1B in prostate cancer and its correlation with poor prognosis ^[31,32]^, our findings unveiled a novel regulatory mechanism of CDKN1B downregulation in cancer and a potential target for future therapeutics development.

Our previous study indicated PUM1 might regulate other cell cycle targets in mice ^[15, 16]^. *PUM1* overexpression in cancer could elicit the growth-promoting effect via repression of targets other than CDKN1B. While other targets of PUM1 in prostate cancer are the subject of the future study, we have found that PUM1-CDKN1b repression appeared to be robust and general among several cell cycle targets in different cancer cells we examined and this axis represented an important regulatory mechanism for cancer cell proliferation. Although the amplification of *PUM1* and *PUM2* locus was detected in a quarter of neuroendocrine prostate cancers ^[33]^, PUM alleles’ gain was not associated with elevated mRNA expression. The mechanism underlying *PUM1* overexpression in prostate cancers is also a subject of future study.

It has been reported that 3’UTR of many key regulators of cancers tended to shorten or lost completely during the tumorigenesis ^[34]^, suggesting that dysregulation in posttranscriptional gene expression may be a common process that contributes to the pathogenesis of neoplasms. Our study of PUM1-mediated translational regulation of cell cycle regulators via binding to their 3’UTR demonstrated an example for such post-transcriptional regulation in cancer and argues for the importance of studying PUM1-CDKN1B regulatory axis in other cancers where the loss of CDKN1B protein expression is associated with tumorigenesis and is a good prognostic marker for cancer survival.

Our findings showed that PUM1 is a pro-proliferative factor in normal and neoplastic cells. It contributed to the emerging concept that post-transcription regulation represents a fundamental regulatory mechanism of cell growth. Further studies on the targets of PUM proteins and PUM protein expression regulation are required to fully elucidate their roles during tumorigenesis. The confirmation that PUM1 is essential for cell cycle progression and prostate tumorigenesis supports the potential role of PUM1 as a therapeutic target in PCa. Further examination of the PUM1-CDKN1B regulatory axis in diverse cancers could help understand how broadly this translational control contributes to tumorigenesis.

## Data Availability

The data used to support the findings of this study are available from the corresponding author upon request.

## Acknowledgments

We would like to thank Dr. Yuan Ji at the University of Chicago for advice and discussion on the analysis of TCGA datasets and his lab for assistance. We appreciate comments from Drs. Jindan Yu and Takeshi Kurita as well as anonymous reviewers. This work was supported by the National Science Foundation of China (31771652 and 81270737).

---

**Supplemental Table I.**
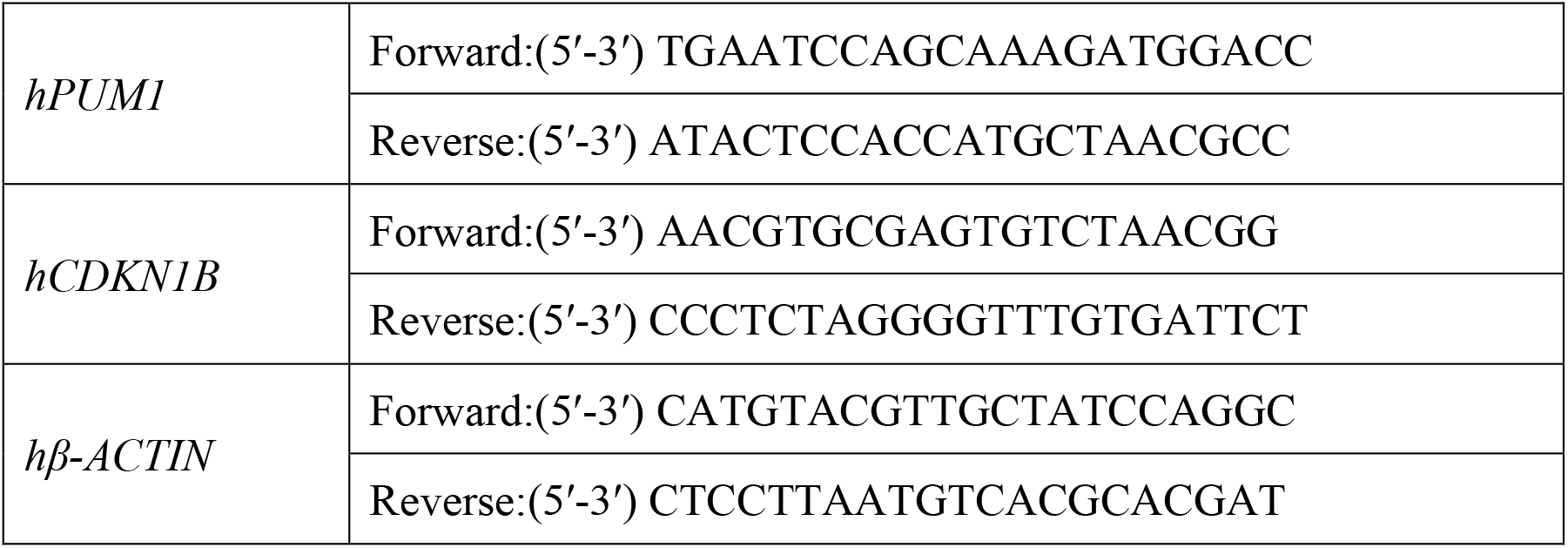

**Supplemental Table II.** Clinical demographics of the prostate cancer cohort (n=20)

|  |  |
| --- | --- |
| Number of patients | 20 |
| Age (y) |  |
| Mean ± SD | 70.5 ± 4.59 |
| Median | 68 |
| Range | 61 - 76 |
| Preoperative PSA (ng/ml) |  |
| Mean ± SD | 12.33 ± 5.42 |
| Median | 11.9 |
| Range | 5.7 – 24.5 |
| Pathological data |  |
| Stage |  |
| pT2 | 15 |
| pT3 | 9 |
| Gleason score |  |
| 7 | 16 |
| >7 | 4 |

